# Impact of Early Interprofessional Involvement with a Critical Care Pharmacist on Patient Outcomes in the Intensive Care Unit

**DOI:** 10.64898/2026.08.25.26361345

**Authors:** Kelli R. Henry, Brooke A. Smith, Devin N. Holden, Susan E. Smith, Mojdeh S. Heavner, Zhetao Chen, Xianyan Chen, John W. Devlin, David J. Murphy, Greg S. Martin, Marisha Burden, Brian Murray, Optimizing Pharmacist Team-Integration for ICU Patient Management (OPTIM) Investigator Team, Andrea Sikora

## Abstract

**Background:** While critical care pharmacists (CCPs) are broadly associated with improvements in outcomes for critically ill patients, operationalizing staffing in the intensive care unit (ICU) requires further study. The purpose of this evaluation was to determine the relationship of a CCP on interprofessional rounds for weekday admissions of ICU patients on patient-centered outcomes.

**Methods:** This post-hoc analysis of the Optimizing Pharmacist-Team Integration for ICU Patient Management (OPTIM) study included adults admitted to an ICU on a weekday in the multicenter observational study. The primary outcome was in-hospital mortality. The primary exposure was level of comprehensive medication management (CMM) during the first 24 hours of ICU stay. A secondary exposure was pharmacist-to-patient ratio. Multivariable generalized estimating equations (GEE) were used to estimate associations between mortality and patient, ICU, and institution variables. Fine-Gray sub-distribution hazards regression estimated hazard of discharge alive (HDA) from the ICU and hospital and hazard of extubation alive.

**Results:** 21,835 patients met inclusion criteria, and 76.1% of patients had CMM delivered on interprofessional rounds. Patients who had no CMM on the first ICU day had an increased risk of mortality of 23% (Odds Ratio (OR) 1.23, 95% Confidence Interval (CI) 1.04-1.46, p=0.02) compared to those who received CMM on interprofessional rounds. Patients with no CMM also had decreased HDA from the ICU and hospital and decreased hazard of extubation alive. No difference was seen in any outcomes when comparing other levels of CMM (CMM delivered outside of interprofessional rounds or abbreviated CMM) compared to CMM delivered on rounds.

**Conclusions:** Absence of pharmacist CMM on the first day of ICU stay for patients with weekday admission was associated with an increased risk of in-hospital mortality, but no difference was seen across levels of CMM: this signal supports further investigation in prospective analysis.

**Key Points:** *Question:* Do ICU patients who do not receive comprehensive medication management (CMM) from a critical care pharmacist (CCP) on interprofessional rounds on the first day have a relationship with worse outcomes?

*Finding:* Patients not receiving CMM had increased mortality and length of stay, but no difference in outcomes was observed in patients receiving partial CMM or CMM delivered outside of interprofessional rounds.

*Meaning:* CMM from a CCP is associated with improved patient outcomes compared to those not receiving CMM, but further research is needed to clarify optimal delivery of CMM and the impact of CCP participation on interprofessional rounds.

## Background

Recent Consensus Recommendations endorsed by five professional organizations including the Society of Critical Care Medicine state that every critically ill patient admitted to an intensive care unit (ICU) requires the care of a critical care pharmacist (CCP).(1) This recommendation is based on a rich body of evidence regarding the importance of comprehensive medication management (CMM) and particularly the attendance of the CCP on interprofessional rounds: including a meta-analysis that showed an association of almost 20% lower odds of mortality with pharmacist attendance and 70% lower adverse drug events (ADEs).(1–17)

The first 24 hours of admission to the ICU is often considered a critical period for patients:1 in 3 deaths in the ICU occur in the first 24 hours, the highest healthcare costs are associated with the first day of admission, and notably, the number of pharmacist medication interventions has been shown to be highest on day 1 of admission.(18–20) However, due to staffing constraints, a CCP may not be present on interprofessional rounds for a variety of reasons, including if they are providing CMM services to multiple rounding teams who conduct interprofessional rounds concurrently, if they are pulled away from rounds due to an emergent situation or need for operational support, or if the shift times required by the Pharmacy Department do not overlap with interprofessional rounding times. Given that the first day of admission is an important window of opportunity to influence outcomes and CCPs on rounds are associated with improved outcomes, we hypothesized that ICU patients who do not receive CMM from a CCP on interprofessional rounds on the first day would have a relationship to worse outcomes.

Understanding the role of individual profession staffing in the complex context of ICU care is challenging to untangle, particularly without resource-intensive prospective data. The purpose of this post-hoc analysis of the **Optimizing Pharmacist Team-Integration for ICU Patient Management** (OPTIM) study was to determine the relationship of having CCP coverage on the first day of a weekday ICU admission on patient mortality and other clinical outcomes. The findings of this study may have potential to inform future ICU workforce study designs and related quality improvement initiatives.

## Methods

### Study Design and Oversight

This study was a post-hoc analysis of a nested cohort of adult patient data from the OPTIM study.(21, 22) OPTIM was a multicenter prospective observational study evaluating the impact of pharmacist workload on patient-centered outcomes, with a protocol and main results previously published.(21–23) The University of Georgia Institutional Review Board (IRB) determined both the original OPTIM study and this post-hoc analysis to be exempt (PROJECT00011467, April 11, 2023). All institutions completed a data use agreement and received IRB approval prior to participation. Data collection used REDCap (Research Electronic Data Capture) hosted by the University of Georgia.(24, 25) Study procedures adhered to the ethical principles outlined in the Helsinki Declaration of 1975.(26) The study design and reporting followed the Strengthening the Reporting of Observational Studies in Epidemiology (STROBE) guidelines for cross-sectional studies (see **Supplementary Appendix)**.(27)

### Study Population

OPTIM included a total of 64 institutions, 33,464 patients, and 213 pharmacists.(22) This post-hoc analysis included patients who were at least 18 years old, had an ICU length of stay (LOS) of at least 24 hours, and were admitted on a weekday (Monday through Friday). Patients were excluded if a transition to comfort measures occurred within the first 24 hours of ICU stay or if they were admitted on a weekend (Saturday or Sunday) (**Figure 1**). Some patients were admitted to a pediatric ICU type; however, all included patients were at least 18 years old.

**Figure 1.**
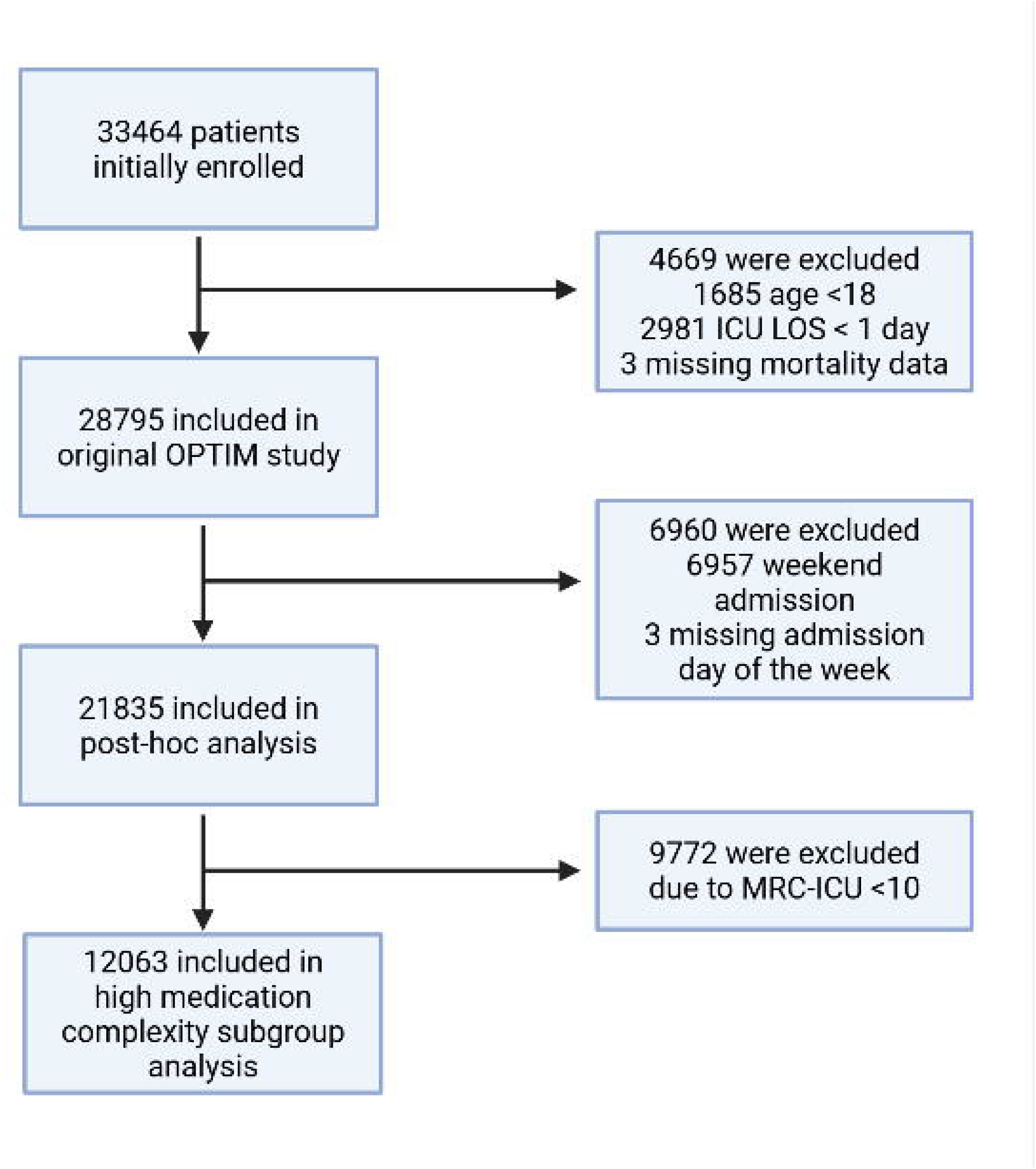
Consort Diagram. ICU: intensive care unit, LOS: length of stay, MRC-ICU: medication regimen complexity-intensive care unit score, OPTIM: Optimizing Pharmacist-Team Integration for ICU Patient Management

### Data Collection and Study Variables

Briefly, prospective data collection occurred from August 2023 to August 2024, with retrospective data collection completed in January 2025.(22) Data collection included 100 days of prospective pharmacist workload data, including the patient-specific pharmacist-to-patient ratio, whether the covering pharmacist performed CMM every day of ICU stay, and whether the covering pharmacist participated in interprofessional rounds every day of ICU stay. Severity of illness variables, including the Sequential Organ Failure Assessment (SOFA) score and the Medication-Regimen Complexity – Intensive Care Unit (MRC-ICU) score were also collected on the first day of ICU admission. (28, 29) Detailed methods are provided in the main analysis, including handling of outside hospital transfers, ICU transfers, and handling of data from non-OPTIM-participating pharmacists.(22)

### Outcomes, Exposures, and Covariates

The primary outcome of this study was in-hospital mortality. Secondary outcomes included ICU hazard of discharge alive (HDA), hospital HDA, and ventilator-free days (VFDs). Fine-Gray Competing Risks Models were used to examine factors associated with the HDA from the ICU and the hazard of VFDs, with mortality treated as a competing event for both outcomes.

The primary exposure of this post-hoc analysis was ICU pharmacist coverage during the first 24 hours of ICU stay. This variable was divided into four categories describing the varying levels of care that a CCP may provide, depending on their staffing structure (**Figure 2**). Standard of care was defined as CMM delivered on interprofessional rounds, defined as the pharmacist attended interprofessional rounds and verbally provided CMM (including review of medications and recommendations) after a full profile review of the patient. This is the gold standard recommended by multiple professional organizations. (1) The second level of care was CMM delivered not on interprofessional rounds, defined as the pharmacist provided CMM by reviewing medications and providing recommendations after a full profile review of the patient but did not attend interprofessional rounds. The third level of care was abbreviated CMM, which included brief medication review but did not include full patient review (e.g. progress notes, results) and recommendations were provided outside of rounds. The ‘final’ (fourth) level of care was CMM not delivered (absence of CMM), meaning that the patient received no medication review from a pharmacist outside of pharmacokinetic monitoring and prospective medication order verification. Prospective order verification includes comprehensive review of the medication at the time of ordering, but does not include followup after initial approval, such as renal dose adjustment, laboratory monitoring, etc. The secondary exposure was pharmacist-to-patient ratio during the first 24 hours of ICU stay. CMM, pharmacist-to-patient ratio, and other CCP workload variables were prospectively recorded by CCPs for each patient.

**Figure 2.**
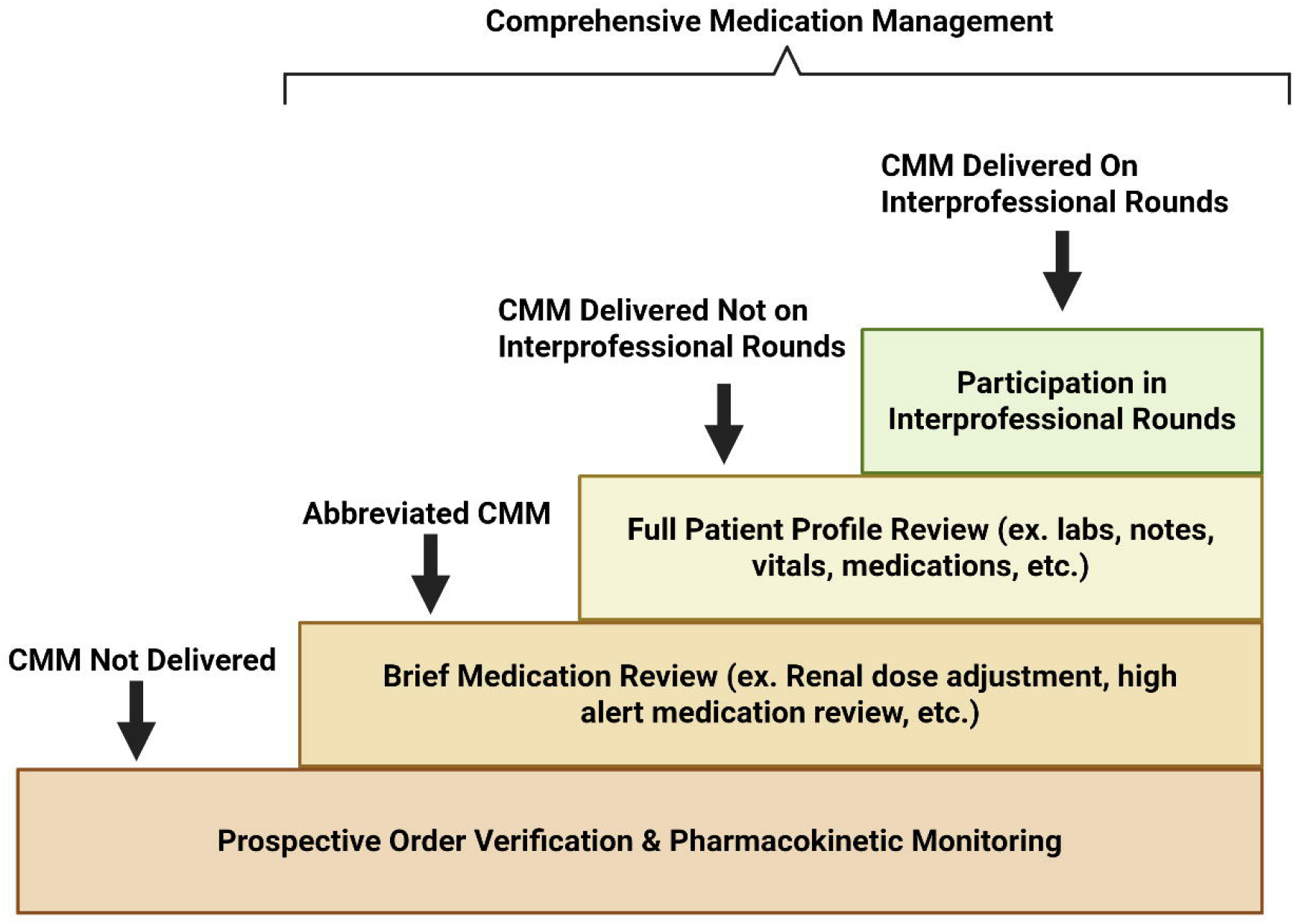
Levels of Comprehensive Medication Management (CMM) This figure portrays the levels of CMM as different critical care pharmacist tasks are added with each increasing level.

Covariates included age, sex, the SOFA and MRC-ICU score during first 24 hours of ICU stay, ICU type, ICU admission day of the week, hospital type, nurse-to-patient ratio on the first day of ICU stay, the proportion of teams the pharmacist rounded with among the total ICU medical teams assigned to the pharmacist on the first day of ICU stay, medical team composition on the first day of ICU stay, presence of a pharmacy trainee on the first day of ICU stay, and institutional Center for Medicare & Medicaid Services (CMS) hospital quality star rating. These covariates were drawn from the primary OPTIM analysis.

### Subgroup Analysis-High Medication Complexity

To evaluate the effect of CMM provided on interprofessional rounds in more medically complex patients, a subgroup analysis of patients with a MRC-ICU score ≥10 in the first 24 hours of admission was completed. Outcomes, exposure variables, and covariates remained the same as in the primary analysis.

### Statistical Analysis

Descriptive statistics, including mean, interquartile range (IQR), counts, and percentages were performed. Missing covariate data were handled using multiple imputation by chained equations, generating 10 imputed datasets under the missing-at-random assumption. Imputation models were selected according to variable type and included predictive mean matching for continuous variables, logistic regression for binary variables, and multinomial logistic regression for categorical variables. Regression models were fitted separately within each imputed dataset, and parameter estimates and standard errors were combined using Rubin’s rules. All statistical tests were two-sided, and statistical significance was defined as p < 0.05. Analyses were performed using R version 4.5.2 (R Foundation for Statistical Computing, Vienna, Austria).

The association between ICU pharmacist coverage during the first 24 hours of ICU stay and in-hospital mortality was evaluated using a population-averaged logistic regression model estimated with generalized estimating equations (GEE), incorporating a logit link, an exchangeable working correlation structure, and robust (sandwich) standard errors. Clustering was specified at the hospital level to account for correlation among patients treated within the same institution. CMM delivered on interprofessional rounds served as the reference category for comparisons across levels of pharmacist coverage. Odds ratios (ORs) with 95% confidence intervals (CI) were reported. All prespecified patient-, ICU-, and institution-level covariates were included in the multivariable model.

Secondary outcomes, including ICU HDA, hospital HDA, and VFDs, were analyzed using Fine-Gray proportional subdistribution hazards models with discharge alive treated as the event of interest and death treated as a competing event. Subdistribution hazard ratios with 95% CIs were reported. Models adjusted for the same covariates included in the mortality analysis and accounted for clustering at the hospital level using robust variance estimation.

A prespecified sensitivity analysis was conducted among patients with a first day MRC-ICU score of ≥10. Subgroup eligibility was determined from the observed data prior to imputation, and the same patient subset was retained across all imputed datasets. The primary and secondary analyses were then repeated using the identical modeling strategy.

## Results

### Baseline characteristics

A total of 33,464 patients were enrolled in OPTIM. After excluding patients if a transition to comfort measures occurred within the first 24 hours of ICU stay or if they were admitted on a weekend (Saturday or Sunday), 21,835 patients were included in this analysis (**Figure 1**). A total of 64 institutions participated, including 62 institutions from the United States, 1 institution from Jordan, and 1 from Saudi Arabia (**Table 1**). The majority (60.9%) of institutions were academic medical centers, and pharmacists covered an average of 2.1 teams per weekday. A total of 216 pharmacists collected data, with the majority having a board certification in critical care pharmacy (71.8%) and completing some form of postgraduate training (91.1%) (**Table 1**).

Patients had an average SOFA score of 5.2 and MRC-ICU score of 11.4 (**Table 2**). Most patients had CMM delivered by a CCP on interprofessional rounds on their first ICU day (76.1%), with 1321 (6.1%) of patients not receiving any CMM on the first day of ICU admission. The average pharmacist-to-patient ratio on the first day of ICU admission was 1:17.7. Patients who received CMM delivered on interprofessional rounds had the lowest pharmacist-to-patient ratios on the first day of admission (mean 1:15.9) compared to patients who received lower levels of CMM. Additional institutional and pharmacist demographics are provided in **Table 1**, and patient demographics and outcomes are provided in **Table 2**. The **Supplementary Appendix eTable 1** provides the patient demographics and outcomes split between alive and deceased patients.

### Primary Outcome

A total of 3,105 patients (14.2%) experienced in-hospital mortality. A multivariable GEE analysis of in-hospital mortality found that the primary exposure variable (ICU pharmacist coverage during the first 24 hours of ICU stay) was associated with increased mortality when comparing patients who did not receive CMM to those who received CMM by a CCP on interprofessional rounds (OR [95% CI]: 1.23 [1.04-1.46]; p = 0.02) (**Table 3**). There was no difference in risk of in-hospital mortality when comparing CMM delivered on interprofessional rounds to CMM delivered outside of interprofessional rounds or abbreviated CMM (**Table 3**). The secondary exposure variable of pharmacist-to-patient ratio was also not associated with increased mortality. Other variables that were associated with in-hospital mortality included age, SOFA score, MRC-ICU score, ICU type, medical team coverage, and nurse-to-patient ratio.**Figure 3** shows the adjusted predicted in-hospital mortality by ICU pharmacist coverage during the first 24 hours of ICU stay.

**Figure 3.**
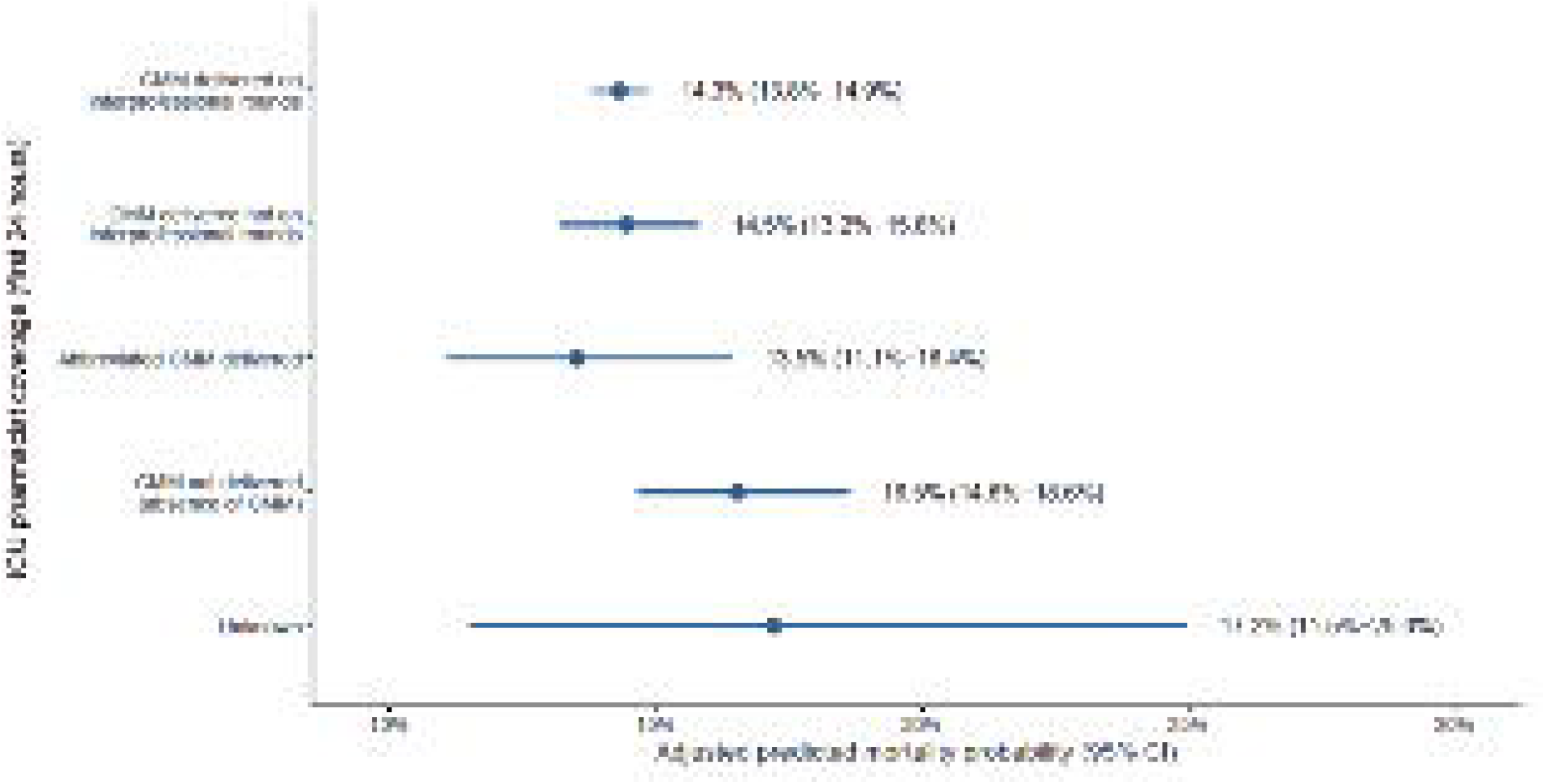
Adjusted Predicted In-Hospital Mortality by ICU Pharmacist Coverage During the First 24 Hours of ICU Stay. CI: confidence interval, CMM: comprehensive medication management, ICU: intensive care unit Note: In-hospital mortality occurred in 14.2% of the cohort.

### Secondary Outcomes

In the Fine-Gray subdistribution hazards regression, no CMM was associated with a significantly lower HDA from the ICU compared to CMM delivered on interprofessional rounds (adjusted hazard ratio (aHR) [95% CI]: 0.78 [0.71–0.85] p = <0.001), a significantly lower HDA from the hospital (aHR [95% CI]: 0.81 [0.73–0.91], p = <0.001), and a significantly lower hazard of extubation alive (aHR [95% CI]: 0.88 [0.82–0.94], p = <0.001) (**Table 4**). There was no difference in other secondary outcomes when comparing other levels of CMM with CMM delivered on interprofessional rounds.

### Subgroup Analysis High Medication Complexity

In the subgroup analysis including only patients with a MRC-ICU score ≥10, 12,063 patients were included. A total of 2,264 patients (18.7%) experienced in-hospital mortality, and the average MRC-ICU score was 16.2 (**Supplementary Appendix eTable 1**). The majority of patients received CMM delivered on interprofessional rounds (76.7%) with 6.1% of patients not receiving CMM on the first day of ICU stay. A mortality GEE analysis did not show an association with ICU pharmacist coverage during the first 24 hours of ICU stay at mortality (**Supplementary Appendix eTable 2**). Age, SOFA score, institution type, ICU type, medical team coverage, and nurse-to-patient ratio had significant associations with risk of in-hospital mortality. Analysis of secondary outcomes (ICU and hospital LOS and VFDs) showed a decreased HDA from the hospital and ICU (aHR 0.83 and 0.75) and decreased hazard of extubation alive (aHR 0.89) for patients who did not receive CMM compared to those who received CMM by a CCP on interprofessional rounds (**Supplementary Appendix eTable 3**). There was no difference in outcomes for patients across levels of CMM during their first 24 hours of ICU stay (**Supplementary Appendix eTable 3**).

## Discussion

This retrospective analysis of the OPTIM study is the first of its kind to evaluate the presence of a pharmacist on interprofessional rounds for the first day of ICU admission. OPTIM is the largest, multi-center evaluation to capture this level of granularity for pharmacist staffing in the context of both patient illness severity and the interprofessional team. This post-hoc analysis explored the effects of various levels of CMM during the first day of ICU admission on ICU and hospital LOS, VFD, and in-hospital mortality after accounting for severity of illness and institutional characteristics. We observed that compared to patients who received CMM from a CCP during interprofessional rounds, patients who did not receive CMM had increased risk of mortality (OR [95% CI]: 1.23 [1.04-1.46], p=0.02) as well as decreased HDA from the hospital and ICU and decreased hazard of extubation alive (aHR 0.81, 0.78, and 0.88, respectively). However, no difference was observed in patient outcomes when comparing CMM on interprofessional rounds to CMM delivered outside of interprofessional rounds or abbreviated CMM. In the analysis of patients with high medication complexity, measured through the MRC-ICU score, there was no difference in mortality risk, but patients who did not receive CMM had a decreased HDA from the hospital and ICU and decreased hazard of extubation alive. While we anticipated to see a difference in CMM delivered on rounds versus outside of rounds specifically in patients with high medication complexity, this analysis was likely underpowered to detect a difference.

These results align with another analysis of OPTIM and have mechanistic plausibility based on other evaluations of CCP workload.(2, 7, 8, 12, 13, 22, 29) The advantage of the present analysis was to focus solely on weekday admissions over any day as global ICU staffing practices of all healthcare workers are often condensed on weekends in a way that confounds potential relationships. While 13.4% of institutions included in OPTIM provided CCP rounding on the weekends, other ICU team members may have higher workloads or decreased provision of services; thus, we included only weekday admissions to reduce this bias. However, there is the possibility that this introduced other bias, as more acutely ill patients may have been admitted on a weekend, as well as difference in other patient characteristics, such as an increased proportion of surgical patients as most elective and non-emergent surgeries may be scheduled for weekdays. Holidays occurring on weekdays were not accounted for, as many institutions did not provide specific dates to maintain patient confidentiality, and holidays were unable to be identified within the dataset. While it may be theorized that CCP attendance on interprofessional rounds may improve patient outcomes, notable gaps still exist in the literature, including if attendance on rounds must happen every day or just some days, if patients with a higher severity of illness are more impacted from CCP presence on rounds, and if the pharmacist-to-patient ratio impacts the ability of the CCP to provide quality CMM on rounds. From OPTIM, it was observed that when compared with no pharmacist, some level of CCP review led to decreased mortality and ICU LOS.(22) A subgroup analysis conducted in OPTIM included a first day analysis using variables associated with the first day of ICU stay, as opposed to averaged across the ICU stay. In this subgroup, there was no association between ICU pharmacist coverage on day 1 of admission with patient mortality. However, this analysis included patients admitted on any day of the week. Studies have shown that patients have worsened outcomes, including mortality if admitted on a weekend due to alterations in staffing of all healthcare providers, including pharmacists, and potential lower provision of resources, including access to surgery, dialysis, procedures, etc.(30–32) This finding is also supported by an evaluation of 10,441 ICU patients that looked to connect pharmacist CMM activity to mortality that observed that most CMM activity occurred early in the ICU admission.(12)

While CMM provided through CCP participation on interprofessional rounds is considered the gold standard, there are logistical barriers that often preclude patients receiving CMM.(1) This reality was observed in the OPTIM study which had 32% of patients who received CMM provided outside of rounds, abbreviated CMM, or no CMM on the first day of ICU admission.(22) These practice variations continue due to equipoise on the best way to logistically deploy pharmacists delivering CMM in the ICU. While pharmacokinetic monitoring, renal dose adjustments, and basic drug information services can be successfully managed regardless of location, the physical presence on interprofessional rounds does facilitate more nuanced discussions with the care team and may result in recommendations that would not have been made otherwise, as new or undocumented information may be revealed in these rich discussions. A study evaluating the effect of an in-person CCP on interprofessional rounds compared to a virtual CCP found that the in-person CCP had significantly more recommendations implemented compared to a virtual CCP.(33) Additionally, pharmacist recommendations provided on interprofessional rounds are evaluated from the perspective of all interprofessional team members for safety, efficacy, and logistics; recommendations communicated outside of interprofessional rounds may not receive the same level of comprehensive evaluation and input from all team members. Further, time to intervention may be decreased as all stakeholders are present for the discussion. Addition of pharmacists on a sepsis response team was shown to have significant improvement in adherence to a one-hour sepsis bundle and decreased time to administration of antibiotics.(34) These findings support other observations of team dynamics in the ICU, which tend to be improved by synchronous communication practices.(35–38)

This study focused on provision of CMM through interprofessional rounds; however, it is important to note that the pharmacist-to-patient ratio within this study aligns with other studies of pharmacist to patient ratio, where increased ratios resulted in poorer outcomes. An institution that capped their pharmacist-to-patient ratio at 1:18 in critical care settings saw a reduction in 30-day readmissions for vulnerable groups.(39) Additionally, a survey found that institutions that met their quality metrics had pharmacist ratios of at least 1:23.(40) Additionally, various committees and organizations have recommended a ratio of 1:15.(1, 41, 42)

While this study did not show a difference in patient outcomes for those who received CMM on interprofessional rounds compared to those who received CMM outside of rounds or partial CMM, there are some key limitations and considerations for applicability of these results. Foremost, this evaluation was a retrospective analysis, which cannot rule out confounding, of a study originally designed to answer a related, yet different, question. Second, while this evaluation specifically focused on the first day of ICU admission, CMM provided throughout the ICU stay likely meaningfully impacts patient outcomes. However, analysis of that relationship at a daily level provides challenges given the heterogeneity of care (e.g., patients may receive CMM on interprofessional rounds one day, but only partial CMM the next day, and no CMM on the last day). Since the original OPTIM study did not collect daily severity of illness variables (such as SOFA score) or daily CCP interventions, it would be challenging to assess the daily provision of CMM without additional context. Future studies could implement more daily data collection to assess the variability in interventions with various staffing models as well as account for fluctuating patient condition. Additionally, while this study tried to account for confounding variables related to variations in staffing models of other ICU professionals (physicians, nurses, physical therapists, etc.) on the weekends by excluding patients who were admitted on a weekend, it is possible that some patients in this analysis, while not admitted on a weekend, were still in the ICU on a weekend (e.g., admitted on a Wednesday, but their ICU stay was Wednesday to Monday), so patients with longer lengths of stay may have received suboptimal care due to variations in weekend staffing from all ICU professions that were not accounted for in this analysis. Additionally, emergency department boarding times were not included, and patients may have received CMM from an emergency medicine pharmacist prior to admission to the ICU. We also chose to exclude patients who transitioned to comfort care or died within the first 24 hours of ICU admission, as we expected that those patients may not have had the opportunity to benefit from CMM provided by a CCP. Several studies have suggested that suboptimal care may occur on weekends; patients admitted on a weekend in OPTIM had an overall mortality rate of 17.2%, compared to the rate of 14.2% seen in the patients included in this analysis.(30–32, 43) Many institutions regularly expect CCPs to cover a large number of patients on the weekends; however, this analysis had an average pharmacist-to-patient ratio of 1:17.7. Therefore, it cannot be extrapolated that provision of CMM can be successfully conducted outside of rounds on the weekends, given that the average pharmacist-to-patient ratio in this study was much lower than what may be expected of a CCP on a weekend, given current staffing models. This analysis sets the stage for future prospective analyses that can more definitely answer these questions.

## Conclusion

OPTIM is the largest cohort study to date examining the effect of CMM on patient outcomes. In this analysis, no CMM on day 1 of ICU admission resulted in significantly higher risk of mortality and decreased HDA, but there was no difference seen between CMM delivered on interprofessional rounds and CMM delivered outside of interprofessional rounds or abbreviated CMM. However, given its retrospective nature and a lack of granular daily data including workload data from other ICU clinicians, this post-hoc analysis may be unable to truly parse the impact of CMM provided on interprofessional rounds on patient outcomes. A prospective study taking into account daily patient severity, ICU and hospital type, ICU clinician workload, and CMM is necessary to determine the optimal mechanism for provision of CMM.

## IRB Approval Information

The protocol was reviewed and approved by the Institutional Review Board at the University of Georgia (April 11, 2023).

## Data Availability

All data produced in the present study are available upon reasonable request to the authors.

## Acknowledgement

All authors contributed to the research and manuscript and meet ICMJE authorship criteria.

