## Supplemental Appendix for "Impact of Early Interprofessional Involvement with a Critical Care Pharmacist on Patient Outcomes in the Intensive Care Unit"

Jessica A. Ward, PharmD, Cleveland Clinic, Cleveland, OH

Nathaniel B. Wayne, PharmD, Wellstar MCG Health, University of Georgia College of Pharmacy, Augusta, GA

Lori Wetmore, PharmD, IU Health, Indianapolis, IN

Jessica A. Whitten, PharmD, Eskenazi Health, Indianapolis, IN

Alexandra M. Wiegand, PharmD, UK HealthCare, Lexington, KY

Sarah K. Williford, PharmD, University of North Carolina Health, Chapel Hill, NC

Sharon Wilson, PharmD, BS, University of Maryland Medical Center, Baltimore, MD

Kevin M. Wohlfarth, PharmD, ProMedica Toledo Hospital/Russel J. Ebeid Children's Hospital, Toledo, OH

Douglas R. Wylie, PharmD, Chiesi USA, Cary, NC

Siu Yan A. Yeung, PharmD, University of Maryland Medical Center, Baltimore, MD

Connie H. Yoon, PharmD, OhioHealth Riverside Methodist Hospital, Columbus, OH

### List of acknowledgements

REDCap Consortium at Vanderbilt

Pharmaceutical Research Computing (PRC) center at University of Maryland School of Pharmacy

Society of Critical Care Medicine (SCCM)

American College of Clinical Pharmacy (ACCP)

Sarah A.H. Adams

Diana Aguilar

Ghadah Alajmi

Muhammad Y. Akbik

May Alanazi

Julia Alexander

Emily Austin

Mark Awad

Erin Beauclair

Cesar Bejarano-Garcia

Andrew T. Bennett

Jessica M. Biggs

Mary Blair

Kaitlin M. Blotske

Christopher Bollinger

James Braun

Nicholas Bravo

Garrett Brown

Jennifer Bui

Joshua Campbell

Carlette Cavenaugh

Michael E. Chao

Aaron Chase

Genevieve Cheung

Sarah Chiu

Patrick Costello

Aubrey A. Defayette

Samantha Delibert

Sam Dewitt

Megan Dorsey

Courtney Feagin

Shelby Fideler

Samantha L. Gauthier

Emily George

Kristen Giles

Alexis Glenn

Renae Gozelski

Aileen Gregorio-Corallo

Liana Ha

Ariel H. Haber

Rachel Hall

Kaylee Hall

Andrea Hankins

Brennan Herrmann

Gresham Hindman

Elizabeth Hodges

Cory Johnson

Sara R. Jones

Kevin Josey

Lama Kanawati

Nadine Kanyana

Jana L. Kelly

Tara Kennell

Alexa Luboff

Isabel Mangaoang

Olivia Marchionda

Carolyn Martz

Taylor McCart

Jamie McCarthy

Hannah McMurrin

Stephanie Millan

Emily Miller

James T. Miller

Makenna Moll

Peter Moran

Rebecca Morgan

Amoreena Most

Zach Muller

Nicole Newton

Kelly T. Nguyen

Christian Nicolosi

Kristine A. Parbuoni

Akta S. Patel

Justin Petrovic

Inna Perinskaya

Nicole Pfeffer

Kara E. Phillips

Leslie Phillips

Lisa Pickmans

Laura Provost

Marinna Raqueno

Paige E. Reese

Jenna Schwartz

Janet Shin

Shaleen Singh

Maya Smith

Alyssa Sonchaiwanich

Jillian K. Songstad

Kelcy Sorsensen

Inderpal Srai

Gillian Steiger

Donna Steinbacher

Samori Swygert

Kari Taggart

Farrah Tavakoli

Dakota Taylor

Melissa Thompson Bastin

Kailee Toews

Meghna Vallabh

Storm A. Van Wey

Noelle Vo

Todd Walroth

Hailey Wang

Brian Watson

Ashley Wischmeyer

Laura Witt

Jae H. Yook

Shelby Young

Denisse Garcia Zavala

Kara Zacholski

Qingrong Laura Zhang

### Reporting of Observational Studies in Epidemiology (STROBE) Checklist

|  | Item No | Recommendation | Page No |
| --- | --- | --- | --- |
| **Title and abstract** | 1 | (*a*) Indicate the study’s design with a commonly used term in the title or the abstract | 1-4 |
|  |  | (*b*) Provide in the abstract an informative and balanced summary of what was done and what was found | 4 |
| Introduction | | | |
| Background/rationale | 2 | Explain the scientific background and rationale for the investigation being reported | 5 |
| Objectives | 3 | State specific objectives, including any prespecified hypotheses | 5 |
| Methods | | | |
| Study design | 4 | Present key elements of study design early in the paper | 6 |
| Setting | 5 | Describe the setting, locations, and relevant dates, including periods of recruitment, exposure, follow-up, and data collection | 6 |
| Participants | 6 | (*a*) Give the eligibility criteria, and the sources and methods of selection of participants. Describe methods of follow-up | 6 |
|  |  | (*b*) For matched studies, give matching criteria and number of exposed and unexposed | n/a |
| Variables | 7 | Clearly define all outcomes, exposures, predictors, potential confounders, and effect modifiers. Give diagnostic criteria, if applicable | 6-7 |
| Data sources/ measurement | 8* | For each variable of interest, give sources of data and details of methods of assessment (measurement). Describe comparability of assessment methods if there is more than one group | 6-7 |
| Bias | 9 | Describe any efforts to address potential sources of bias | 7 |
| Study size | 10 | Explain how the study size was arrived at | 7 |
| Quantitative variables | 11 | Explain how quantitative variables were handled in the analyses. If applicable, describe which groupings were chosen and why | 7 |
| Statistical methods | 12 | (*a*) Describe all statistical methods, including those used to control for confounding | 7 |
|  |  | (*b*) Describe any methods used to examine subgroups and interactions | n/a |
|  |  | (*c*) Explain how missing data were addressed | n/a |
|  |  | (*d*) If applicable, explain how loss to follow-up was addressed | n/a |
|  |  | (*e*) Describe any sensitivity analyses | n/a |
| Results | | |  |
| Participants | 13* | (a) Report numbers of individuals at each stage of study—eg numbers potentially eligible, examined for eligibility, confirmed eligible, included in the study, completing follow-up, and analysed | 8, Figure 1 |
|  |  | (b) Give reasons for non-participation at each stage | Figure 1 |
|  |  | (c) Consider use of a flow diagram | Figure 1 |
| Descriptive data | 14* | (a) Give characteristics of study participants (eg demographic, clinical, social) and information on exposures and potential confounders | 8 |
|  |  | (b) Indicate number of participants with missing data for each variable of interest | Tables 1 and 2 |
|  |  | (c) Summarise follow-up time (eg, average and total amount) | 8 |
| Outcome data | 15* | Report numbers of outcome events or summary measures over time | Table 2 |

| Main results | 16 | (*a*) Give unadjusted estimates and, if applicable, confounder-adjusted estimates and their precision (eg, 95% confidence interval). Make clear which confounders were adjusted for and why they were included | 8, Table 3 |
| --- | --- | --- | --- |
|  |  | (*b*) Report category boundaries when continuous variables were categorized | 8 |
|  |  | (*c*) If relevant, consider translating estimates of relative risk into absolute risk for a meaningful time period | 8 |
| Other analyses | 17 | Report other analyses done—eg analyses of subgroups and interactions, and sensitivity analyses | 8-9, Supplemental |
| Discussion | | | |
| Key results | 18 | Summarise key results with reference to study objectives | 10 |
| Limitations | 19 | Discuss limitations of the study, taking into account sources of potential bias or imprecision. Discuss both direction and magnitude of any potential bias | 11 |
| Interpretation | 20 | Give a cautious overall interpretation of results considering objectives, limitations, multiplicity of analyses, results from similar studies, and other relevant evidence | 10-11 |
| Generalisability | 21 | Discuss the generalisability (external validity) of the study results | 10-11 |
| Other information | | | |
| Funding | 22 | Give the source of funding and the role of the funders for the present study and, if applicable, for the original study on which the present article is based | 1 |

*Give information separately for exposed and unexposed groups.

**Note:** An Explanation and Elaboration article discusses each checklist item and gives methodological background and published examples of transparent reporting. The STROBE checklist is best used in conjunction with this article (freely available on the Web sites of PLoS Medicine at http://www.plosmedicine.org/, Annals of Internal Medicine at http://www.annals.org/, and Epidemiology at http://www.epidem.com/). Information on the STROBE Initiative is available at http://www.strobe-statement.org.

### Disclosures

Authors with conflicts of interest are listed below. If an author is not listed, they reported no conflicts of interest.

### eTable 1. Patient demographics, intensity of pharmacist coverage and outcomes

| **Variable** | **Overall**  **(21,835)** | **Deceased**  **(3,105)** | **Alive**  **(18,730)** | **p-value** |
| --- | --- | --- | --- | --- |
| ***Primary Variable*** | | | |  |
| ICU pharmacist coverage 1^st^ 24 hours**‡***  CMM delivered on interprofessional rounds  CMM delivered not on interprofessional rounds  Abbreviated CMM delivered  CMM not delivered (absence of CMM)  Unknown  Missing | 16,605 (76.1)  2,998 (13.7)  810 (3.7)  1,321 (6.1)  95 (0.4)  6 (<0.1) | 2,394 (77.1)  377 (12.1)  100 (3.2)  214 (6.9)  20 (0.6)  0 (0) | 14,211 (75.9)  2,621 (14.0)  710 (3.8)  1,107 (5.9)  75 (0.4)  6 (<0.1) | 0.002 |
| ***Secondary Variable*** | | | | |
| Pharmacist-to-patient ratio (ICU patients)*  Missing | 17.7 (±10.5)  1,148 (5.3) | 17.7 (±11.3)  174 (5.6) | 17.7 (±10.4)  974 (5.2) | 0.12 |
| ***Co-variates*** | | | | |
| Age, years | 61.6 (±16.7) | 65.8 (±15.2) | 60.9 (±16.8) | <0.001 |
| Sex, female | 9,226 (42.3) | 1,299 (41.8) | 7,927 (42.3) | 0.4 |
| SOFA Score*  Missing | 5.2 (±4.1)  25 (0.1) | 8.6 (±4.4)  2 (<0.1) | 4.6 (±3.8)  23 (0.1) | <0.001 |
| MRC-ICU Score*  Missing | 11.4 (±6.6)  2 (<0.1)\ | 14.5 (±7.1)  0 (0) | 10.9 (±6.4)  2 (<0.1) | <0.001 |
| ICU admission day of the week  Monday  Tuesday  Wednesday  Thursday  Friday | 4,634 (21.2)  4,599 (21.1)  4,364 (20.0)  4,386 (20.1)  3,852 (17.6) | 669 (21.5)  640 (20.6)  615 (19.8)  623 (20.1)  558 (18.0) | 3,965 (21.2)  3,959 (21.1)  3,749 (20.0)  3,763 (20.1)  3,294 (17.6) | >0.9 |
| ICU type**†***  Medical  Surgical/Trauma  Surgical  Cardiothoracic  Cardiac  Neurosurgery/Neurology  Mixed Medical-Surgical  Pediatric  Burn  Other  Missing | 6,568 (30.1)  2,757 (12.6)  2,026 (9.3)  2,272 (10.4)  1,964 (9.0)  2,928 (13.4)  3,113 (14.3)  27 (0.1)  141 (0.6)  37 (0.2)  2 (<0.1) | 1,292 (41.6)  295 (9.5)  222 (7.1)  153 (4.9)  310 (10.0)  343 (11.0)  473 (15.2)  1 (0.0)  9 (0.3)  7 (0.2)  0 (0) | 5,276 (28.2)  2,462 (13.1)  1,804 (9.6)  2,119 (11.3)  1,654 (8.8)  2,585 (13.8)  2,640 (14.1)  26 (0.1)  132 (0.7)  30 (0.2)  2 (<0.1) | <0.001 |
| Hospital type  Academic Medical Center  Community - Teaching  Community - Non-teaching  Government/VA/Military | 14,590 (66.8)  5,322 (24.4)  1,797 (8.2)  126 (0.6) | 2,098 (67.6)  768 (24.7)  225 (7.2)  14 (0.5) | 12,492 (66.7)  4,554 (24.3)  1,572 (8.4)  112 (0.6) | 0.12 |
| Medical team coverage*  Attending physician only  Attending physician and APP  Attending physician and medical residents/fellows  Attending physician and medical residents/fellows and APP  Missing | 1,772 (8.2)  5,065 (23.6)  7,178 (33.4)  7,464 (34.8)  356 (1.6) | 261 (8.6)  607 (19.9)  1,118 (36.7)  1,058 (34.8)  61 (2.0) | 1,511 (8.2)  4,458 (24.2)  6,060 (32.9)  6,406 (34.7)  295 (1.6) | <0.001 |
| Hospital CMS star rating  2  3  4  5  Missing | 4,853 (22.8)  11,205 (52.6)  4,922 (23.1)  334 (1.6)  521 (2.4) | 639 (21.7)  1,561 (53.0)  712 (24.2)  36 (1.2)  157 (5.1) | 4,214 (22.9)  9,644 (52.5)  4,210 (22.9)  298 (1.6)  364 (1.9) | 0.1 |
| Pharmacy trainee present*  No  Yes  Missing | 12,388 (57.8)  9,041 (42.2)  406 (1.9) | 1,756 (57.4)  1,304 (42.6)  45 (1.4) | 10,632 (57.9)  7,737 (42.1)  361 (1.9) | 0.6 |
| Nurse-to-patient ratio, 1:X*  ≤1  1-2  2  >2  Missing | 2,432 (12.0)  47 (0.2)  17,620 (86.7)  229 (1.1)  1,507 (6.9) | 503 (17.4)  5 (0.2)  2,367 (81.8)  20 (0.7)  210 (6.8) | 1,929 (11.1)  42 (0.2)  15,253 (87.5)  209 (1.2)  1,297 (6.9) | <0.001 |
| Percent of assigned ICU teams pharmacist rounded with**§***  Missing | 66.2 (34.0)  1,157 (5.3) | 68.6 (33.3)  175 (5.6) | 65.8 (34.1)  982 (5.2) | <0.001 |
| CMM delivered every day of ICU stay | 16,696 (76.5) | 2,207 (71.1) | 14,489 (77.4) | <0.001 |
| Percent days, only attending physician**⁂**  Missing | 8.6 (26.4)  1,326 (6.1) | 8.7 (25.8)  284 (9.1) | 8.6 (26.5)  1,042 (5.6) | 0.2 |
| Required dialysis during ICU stay  No  Yes  Missing | 19,273 (88.3)  2,560 (11.7)  2 (<0.1) | 2,233 (72.0)  870 (28.0)  2 (<0.1) | 17,040 (91.0)  1,690 (9.0)  0 (0) | <0.001 |
| Required mechanical ventilation | 10,733 (49.2) | 2,330 (75.0) | 8,403 (44.9) | <0.001 |
| Mean pharmacist-to-patient ratio (ICU Patients) across hospital stay  Missing | 19.4 (±10.1)  279 (1.3) | 19.2 (±9.8)  29 (0.9) | 19.4 (±10.2)  250 (1.3) | 0.6 |
| Mean pharmacist-to-patient ratio (all patients) across hospital stay  Missing | 27.2 (±22.7)  279 (1.3) | 26.0 (±19.3)  29 (0.9) | 27.4 (±23.2)  250 (1.3) | 0.024 |
| ***Outcomes*** | | | |  |
| In hospital mortality | -- | 3,105 (14.2) | 18,730 (85.8) | -- |
| Hospital LOS, days  Missing | 12.2 (±16.9)  2 (<0.1) | 14.4 (±20.5)  0 (0) | 11.9 (±16.2)  2 (<0.1) | <0.001 |
| ICU LOS, days | 5.7 (±9.7) | 8.7 (±13.4) | 5.2 (±8.9) | <0.001 |
| Duration of mechanical ventilation, days  Missing | 2.3 (±20.5)  1 (<0.1) | 5.2 (±8.5)  0 (0) | 1.8 (±21.9)  1 (<0.1) | <0.001 |

Data presented as mean (±SD) or n (%)

APP: advanced practice provider, CMM: comprehensive medication management, CMS: Center for Medicare & Medicaid Services, ICU: intensive care unit, LOS: length of stay, MRC-ICU: medication regimen complexity- intensive care unit, SOFA: sequential organ failure assessment, VA: Veterans Affairs

Full details of missingness are available in the **Supplementary Appendix.**

*indicates that this is reflective of only the value on the first day of ICU stay

**†**Inclusion criteria specified ≥18 years of age; however, some institutions allowed patients ≥18 years of age in the pediatric ICU

**‡**CMM delivered on interprofessional rounds: the pharmacist attended interprofessional rounds and verbally provided CMM (including review of medications and recommendations); CMM delivered not on interprofessional rounds: the pharmacist provided CMM (reviewed medications and provided recommendations) but did not attend interprofessional rounds; Abbreviated CMM delivered: the pharmacist provided abbreviated CMM which includes brief medication review but does not include full patient review (e.g. progress notes, results) and recommendations were provided outside of rounds; CMM not delivered (absence of CMM): the patient received no medication review from a pharmacist outside of pharmacokinetic monitoring and prospective medication order verification

**§**Percent of assigned ICU teams pharmacist rounded is calculated by total number of ICU medical teams the pharmacist attended and provided CMM on interprofessional rounds for divided by total number of ICU medical teams providing care for patients that the pharmacist was assigned to provide CMM for

### Subgroup Analysis – High Medication Complexity

#### **eTable 2**. Patient demographics, intensity of pharmacist coverage and outcomes for patients with an MRC-ICU ≥ 10

| **Variable** | **Overall**  **(12,063)** | **Deceased**  **(2,264)** | **Alive**  **(9,799)** | **p-value** |
| --- | --- | --- | --- | --- |
| ***Primary Variable*** | | | |  |
| ICU pharmacist coverage 1^st^ 24 hours**‡***  CMM delivered on interprofessional rounds  CMM delivered not on interprofessional rounds  Abbreviated CMM delivered  CMM not delivered (absence of CMM)  Unknown | 9,250 (76.7)  1,581 (13.1)  430 (3.6)  732 (6.1)  66 (0.5) | 1,757 (77.6)  271 (12.0)  74 (3.3)  149 (6.6)  13 (0.6) | 1,757 (77.6)  1,310 (13.4)  356 (3.6)  583 (6.0)  53 (0.5) | 0.3 |
| ***Secondary Variable*** | | | | |
| Pharmacist-to-patient ratio (ICU patients)* | 19.4 (9.6) | 19.0 (9.1) | 19.4 (9.7) | 0.2 |
| ***Co-variates*** | | | | |
| Age, years | 60.2 (16.3) | 64.6 (15.1) | 59.2 (16.4) | <0.001 |
| Sex, female | 4,863 (40.3) | 927 (40.9) | 3,936 (40.2) | 0.6 |
| SOFA Score* | 7.0 (4.1) | 9.8 (4.2) | 6.4 (3.8) | <0.001 |
| MRC-ICU Score* | 16.2 (5.1) | 17.6 (5.7) | 15.8 (4.8) | <0.001 |
| ICU admission day of the week  Monday  Tuesday  Wednesday  Thursday  Friday | 2,527 (20.9)  2,480 (20.6)  2,441 (20.2)  2,441 (20.2)  2,174 (18.0) | 485 (21.4)  466 (20.6)  455 (20.1)  462 (20.4)  396 (17.5) | 2,042 (20.8)  2,014 (20.6)  1,986 (20.3)  1,979 (20.2)  1,778 (18.1) | >0.9 |
| ICU type**†***  Medical  Surgical/Trauma  Surgical  Cardiothoracic  Cardiac  Neurosurgery/Neurology  Mixed Medical-Surgical  Pediatric  Burn  Other | 3,536 (29.3)  1,531 (12.7)  1,249 (10.4)  1,833 (15.2)  964 (8.0)  1,189 (9.9)  1,648 (13.7)  9 (0.1)  81 (0.7)  22 (0.2) | 913 (40.3)  223 (9.8)  172 (7.6)  124 (5.5)  225 (9.9)  228 (10.1)  364 (16.1)  1 (0.0)  8 (0.4)  6 (0.3) | 2,623 (26.8)  1,308 (13.3)  1,077 (11.0)  1,709 (17.4)  739 (7.5)  961 (9.8)  1,284 (13.1)  8 (0.1)  73 (0.7)  16 (0.2) | <0.001 |
| Hospital type  Academic Medical Center  Community - Teaching  Community - Non-teaching  Government/VA/Military | 8,279 (68.6)  2,766 (22.9)  985 (8.2)  33 (0.3) | 1,521 (67.2)  570 (25.2)  167 (7.4)  6 (0.3) | 6,758 (69.0)  2,196 (22.4)  818 (8.3)  27 (0.3) | 0.028 |
| Medical team coverage*  Attending physician only  Attending physician and APP  Attending physician and medical residents/fellows  Attending physician and medical residents/fellows and APP | 885 (7.5)  2,906 (24.5)  3,945 (33.3)  4,110 (34.7) | 193 (8.7)  451 (20.3)  800 (36.0)  778 (35.0) | 692 (7.2)  2,455 (25.5)  3,145 (32.7)  3,332 (34.6) | <0.001 |
| Hospital CMS star rating  2  3  4  5 | 2,567 (21.8)  5,958 (50.7)  3,024 (25.7)  204 (1.7) | 460 (21.4)  1,121 (52.2)  542 (25.2)  24 (1.1) | 2,107 (21.9)  4,837 (50.4)  2,482 (25.8)  180 (1.9) | 0.06 |
| Pharmacy trainee present* | 5,066 (42.9) | 957 (43.0) | 4,109 (42.8) | 0.9 |
| Nurse-to-patient ratio, 1:X*  ≤1  1-2  2  >2 | 1,754 (15.7)  28 (0.3)  9,298 (83.3)  82 (0.7) | 424 (20.0)  4 (0.2)  1,673 (79.1)  14 (0.7) | 1,330 (14.7)  24 (0.3)  7,625 (84.3)  68 (0.8) | <0.001 |
| Percent of assigned ICU teams pharmacist rounded with**§*** | 66.2 (34.1) | 68.3 (33.6) | 65.7 (34.2) | <0.001 |
| CMM delivered every day of ICU stay | 8,908 (73.8) | 1,607 (71.0) | 7,301 (74.5) | <0.001 |
| Percent days, only attending physician**⁂** | 7.9 (24.9) | 8.9 (25.9) | 7.7 (24.7) | <0.004 |
| Required dialysis during ICU stay | 1,821 (15.1) | 708 (31.3) | 1,113 (11.4) | <0.001 |
| Required mechanical ventilation | 8,924 (74.0) | 1,928 (85.2) | 6,996 (71.4) | <0.001 |
| Pharmacist-to-patient ratio (ICU Patients) averaged across hospital stay | 19.4 (9.6) | 19.0 (9.1) | 19.4 (9.7) | 0.2 |
| Pharmacist-to-patient ratio (all patients) averaged across hospital stay | 26.6 (19.4) | 25.8 (18.1) | 26.8 (19.7) | 0.013 |
| ***Outcomes*** | | | |  |
| Hospital LOS, days | 14.7 (18.8) | 14.8 (20.9) | 14.7 (18.2) | <0.001 |
| ICU LOS, days | 7.2 (11.5) | 9.3 (13.9) | 6.8 (10.8) | <0.001 |
| Duration of mechanical ventilation, days | 3.6 (27.4) | 6.0 (8.9) | 3.0 (30.1) | <0.001 |

Data presented as mean (±SD) or n (%)

APP: advanced practice provider, CMM: comprehensive medication management, CMS: Center for Medicare & Medicaid Services, ICU: intensive care unit, LOS: length of stay, MRC-ICU: medication regimen complexity- intensive care unit, SOFA: sequential organ failure assessment, VA: Veterans Affairs

*indicates that this is reflective of only the value on the first day of ICU stay

**†**Inclusion criteria specified ≥18 years of age; however, some institutions allowed patients ≥18 years of age in the pediatric ICU

**‡**CMM delivered on interprofessional rounds: the pharmacist attended interprofessional rounds and verbally provided CMM (including review of medications and recommendations); CMM delivered not on interprofessional rounds: the pharmacist provided CMM (reviewed medications and provided recommendations) but did not attend interprofessional rounds; Abbreviated CMM delivered: the pharmacist provided abbreviated CMM which includes brief medication review but does not include full patient review (e.g. progress notes, results) and recommendations were provided outside of rounds; CMM not delivered (absence of CMM): the patient received no medication review from a pharmacist outside of pharmacokinetic monitoring and prospective medication order verification

**§**Percent of assigned ICU teams pharmacist rounded is calculated by total number of ICU medical teams the pharmacist attended and provided CMM on interprofessional rounds for divided by total number of ICU medical teams providing care for patients that the pharmacist was assigned to provide CMM for

#### **eTable 3**. Unadjusted and adjusted odds ratios for GEE for primary outcome of mortality

| **Variable** | **Univariable** | | **Multivariable** | |
| --- | --- | --- | --- | --- |
|  | **Odds ratio (CI)** | **P-value** | **Odds Ratio (CI)** | **P-value** |
| ***Primary Variable*** | | | |  |
| ICU pharmacist coverage 1st 24 hours‡* (reference: CMM delivered on interprofessional rounds)  CMM delivered not on interprofessional rounds  Abbreviated CMM delivered  CMM not delivered (absence of CMM)  Unknown | 0.92 (0.78–1.08)  0.96 (0.78–1.18)  1.02 (0.85–1.23)  1.12 (0.47–2.67) | 0.32  0.68  0.8  0.81 | 1.04 (0.89–1.23)  1.05 (0.78–1.41)  1.18 (0.93–1.48)  1.01 (0.44–2.36) | 0.6  0.75  0.17  0.97 |
| ***Secondary Variable*** | | | | |
| Pharmacist-to-patient ratio (ICU patients)* | 1.00 (0.99–1.01) | 0.77 | 1.00 (1.00–1.01) | 0.31 |
| ***Co-variates*** | | | | |
| Age, years | 1.02 (1.02–1.03) | <0.001 | 1.03 (1.02–1.03) | <0.001 |
| Sex, female | 1.02 (0.93–1.11) | 0.74 | 1.00 (0.91–1.11) | 0.94 |
| SOFA Score* | 1.24 (1.22–1.26) | <0.001 | 1.24 (1.17–1.30) | <0.001 |
| MRC-ICU Score* | 1.07 (1.05–1.09) | <0.001 | 1.01 (0.98–1.04) | 0.54 |
| ICU admission day of the week (reference: Monday)  Tuesday  Wednesday  Thursday  Friday | 0.98 (0.85–1.12)  0.96 (0.82–1.13)  0.97 (0.83–1.15)  0.93 (0.80–1.08) | 0.73  0.64  0.76  0.35 | 1.00 (0.86–1.16)  0.94 (0.79–1.12)  1.00 (0.84–1.19)  0.86 (0.73–1.00) | 0.98  0.51  0.96  0.05 |
| ICU type†* (reference: medical ICU)  Surgical/Trauma  Surgical  Cardiothoracic  Cardiac  Neurosurgery/Neurology  Mixed Medical-Surgical  Pediatric  Burn  Other | 0.51 (0.42–0.63)  0.48 (0.34–0.68)  0.22 (0.15–0.33)  0.84 (0.64–1.10)  0.68 (0.56–0.82)  0.78 (0.60–1.02)  0.36 (0.06–2.13)  0.36 (0.21–0.62)  1.00 (0.55–1.82) | <0.001  <0.001  <0.001  0.2  <0.001  0.07  0.26  <0.001  0.99 | 0.75 (0.60–0.95)  0.52 (0.38–0.70)  0.20 (0.13–0.32)  0.95 (0.70–1.29)  1.28 (0.99–1.66)  0.77 (0.61–0.96)  0.98 (0.22–4.42)  0.63 (0.34–1.19)  1.44 (0.86–2.41) | 0.02  <0.001  <0.001  0.75  0.06  0.02  0.98  0.15  0.16 |
| Hospital type (reference: academic medical center)  Community - Teaching  Community - Non-teaching  Government/VA/Military | 1.08 (0.82–1.43)  0.88 (0.63–1.23)  0.93 (0.77–1.12) | 0.57  0.46  0.43 | 0.97 (0.78–1.20)  0.74 (0.56–0.99)  0.79 (0.63–0.98) | 0.76  0.04  0.03 |
| Medical team coverage* (reference: only attending physician)  Attending physician and APP  Attending physician and medical residents/fellows  Attending physician and medical residents/fellows and APP | 0.78 (0.53–1.17)  1.05 (0.71–1.54)  0.94 (0.62–1.43) | 0.23  0.82  0.77 | 0.71 (0.54–0.93)  0.76 (0.56–1.02)  0.80 (0.58–1.11) | 0.01  0.07  0.19 |
| Hospital CMS star rating (reference: 3)  2  4  5 | 0.94 (0.74–1.19)  1.03 (0.78–1.36)  0.68 (0.28–1.63) | 0.58  0.82  0.38 | 0.98 (0.79–1.22)  1.02 (0.80–1.30)  0.64 (0.40–1.04) | 0.86  0.88  0.07 |
| Pharmacy trainee present* |  |  |  |  |
| Nurse-to-patient ratio* (reference: 1:2)  ≤1  1-2  >2 | 1.67 (1.29–2.17)  0.75 (0.45–1.23)  0.83 (0.56–1.25) | <0.001  0.24  0.38 | 1.52 (1.11–2.08)  0.81 (0.56–1.18)  0.90 (0.58–1.42) | 0.009  0.27  0.66 |
| Percent of assigned ICU teams pharmacist rounded with**§*** | 1.00 (1.00–1.00) | 0.15 | 1.00 (1.00–1.00) | 0.17 |
| SOFA-MRC-ICU interaction term | 1.01 (1.01–1.01) | <0.001 | 1.00 (1.00–1.00) | 0.81 |

APP: advanced practice provider, CI: confidence interval, CMM: comprehensive medication management, CMS: Center for Medicare & Medicaid Services, ICU: intensive care unit, LOS: length of stay, MRC-ICU: medication regimen complexity- intensive care unit, SOFA: sequential organ failure assessment, VA: Veterans Affairs

*indicates that this is reflective of only the value on the first day of ICU stay

**†**Inclusion criteria specified ≥18 years of age; however, some institutions allowed patients ≥18 years of age in the pediatric ICU

**‡**CMM delivered on interprofessional rounds: the pharmacist attended interprofessional rounds and verbally provided CMM (including review of medications and recommendations); CMM delivered not on interprofessional rounds: the pharmacist provided CMM (reviewed medications and provided recommendations) but did not attend interprofessional rounds; Abbreviated CMM delivered: the pharmacist provided abbreviated CMM which includes brief medication review but does not include full patient review (e.g. progress notes, results) and recommendations were provided outside of rounds; CMM not delivered (absence of CMM): the patient received no medication review from a pharmacist outside of pharmacokinetic monitoring and prospective medication order verification

**§**Percent of assigned ICU teams pharmacist rounded is calculated by total number of ICU medical teams the pharmacist attended and provided CMM on interprofessional rounds for divided by total number of ICU medical teams providing care for patients that the pharmacist was assigned to provide CMM for

#### **eTable 4**. Fine-Gray Sub-Distribution Hazards Regression for HDA from Hospital and ICU and Hazard of Extubation Alive

|  | **HDA from Hospital** | | **HDA from ICU** | | **Hazard of Extubation Alive** | |
| --- | --- | --- | --- | --- | --- | --- |
|  | **Adjusted Hazard Ratio (95% CI)** | **P-value** | **Adjusted Hazard Ratio (95% CI)** | **P-value** | **Adjusted Hazard Ratio (95% CI)** | **P-value** |
| ***Primary Variable*** | | | |  |  |  |
| ICU pharmacist coverage 1st 24 hours‡* (reference: CMM delivered on interprofessional rounds)  CMM delivered not on interprofessional rounds  Abbreviated CMM delivered  CMM not delivered (absence of CMM)  Unknown | 0.94 (0.87, 1.02)  0.93 (0.80, 1.08)  0.83 (0.74, 0.93)  0.74 (0.54, 1.02) | 0.16  0.33  0.001  0.07 | 1.01 (0.93, 1.09)  1.03 (0.89, 1.19)  0.75 (0.68, 0.84)  0.67 (0.51, 0.88) | 0.85  0.69  <0.001  0.003 | 1.04 (0.96, 1.13)  1.00 (0.88, 1.14)  0.89 (0.81, 0.98)  0.86 (0.68, 1.08) | 0.32  0.94  0.01  0.2 |
| ***Secondary Variable*** | | | | |  |  |
| Pharmacist-to-patient ratio (ICU patients)* | 1.00 (1.00, 1.00) | 0.27 | 1.00 (0.99, 1.00) | 0.05 | 1.00 (1.00, 1.00) | 0.14 |
| ***Co-variates*** | | | | |  |  |
| Age, years | 0.99 (0.99, 0.99) |  | 1.00 (1.00, 1.00) | 0.1 | 0.99 (0.99, 1.00) | <0.001 |
| Sex, female | 0.99 (0.94, 1.03) | 0.59 | 0.97 (0.93, 1.02) | 0.24 | 1.00 (0.96, 1.05) | 0.95 |
| SOFA Score* | 0.90 (0.89, 0.91) | <0.001 | 0.93 (0.92, 0.94) | <0.001 | 0.89 (0.88, 0.90) | <0.001 |
| MRC-ICU Score* | 0.98 (0.98, 0.99) | <0.001 | 0.98 (0.98, 0.99) | <0.001 | 0.98 (0.97, 0.98) | <0.001 |
| ICU admission day of the week (reference: Monday)  Tuesday  Wednesday  Thursday  Friday | 1.01 (0.94, 1.08)  1.05 (0.98, 1.12)  0.99 (0.93, 1.05)  1.05 (0.97, 1.14) | 0.89  0.13  0.71  0.22 | 1.00 (0.94, 1.07)  1.02 (0.97, 1.08)  0.99 (0.94, 1.05)  1.00 (0.93, 1.08) | 0.9  0.41  0.83  0.97 | 1.00 (0.93, 1.07)  1.01 (0.95, 1.08)  0.99 (0.94, 1.06)  1.00 (0.93, 1.08) | 0.92  0.78  0.87  0.92 |
| ICU type†* (reference: medical ICU)  Surgical/Trauma  Surgical  Cardiothoracic  Cardiac  Neurosurgery/Neurology  Mixed Medical-Surgical  Pediatric  Burn  Other | 1.05 (0.90, 1.23)  1.13 (0.96, 1.32)  2.09 (1.75, 2.48)  1.03 (0.87, 1.22)  0.85 (0.74, 0.97)  1.12 (0.98, 1.29)  1.08 (0.55, 2.11)  1.00 (0.59, 1.71)  1.05 (0.48, 2.33) | 0.5  0.14  <0.001  0.73  0.02  0.11  0.82  1  0.9 | 0.89 (0.74, 1.06)  1.16 (0.93, 1.44)  1.45 (1.20, 1.76)  0.97 (0.79, 1.20)  0.73 (0.60, 0.88)  1.28 (1.04, 1.58)  0.89 (0.40, 1.99)  0.72 (0.46, 1.13)  0.65 (0.27, 1.57) | 0.2  0.19  <0.001  0.8  0.001  0.02  0.77  0.15  0.34 | 1.06 (0.95, 1.17)  1.48 (1.27, 1.72)  2.35 (1.99, 2.77)  1.16 (1.00, 1.35)  0.82 (0.73, 0.92)  1.20 (1.04, 1.38)  1.25 (0.84, 1.87)  1.10 (0.67, 1.79)  0.87 (0.45, 1.68) | 0.32  <0.001  <0.001  0.05  <0.001  0.01  0.28  0.71  0.67 |
| Hospital type (reference: academic medical center)  Community - Teaching  Community - Non-teaching  Government/VA/Military | 0.97 (0.86, 1.09)  1.77 (1.50, 2.08)  2.50 (1.50, 4.18) | 0.56  <0.001  <0.001 | 1.07 (0.92, 1.23)  0.75 (0.63, 0.91)  0.68 (0.45, 1.02) | 0.38  0.002  .06 | 0.89 (0.80, 1.00)  1.02 (0.87, 1.19)  1.49 (1.04, 2.15) | 0.05  0.82  0.03 |
| Medical team coverage* (reference: only attending physician)  Attending physician and APP  Attending physician and medical residents/fellows  Attending physician and medical residents/fellows and APP | 1.11 (0.94, 1.32)  1.10 (0.92, 1.31)  1.09 (0.91, 1.32) | 0.2  0.3  0.34 | 1.20 (1.03, 1.40)  1.15 (0.99, 1.34)  1.21 (1.02, 1.43) | 0.02  0.06  0.03 | 1.22 (1.03, 1.44)  1.13 (0.95, 1.33)  1.10 (0.92, 1.32) | 0.02  0.17  0.29 |
| Hospital CMS star rating (reference: 3)  2  4  5 | 0.88 (0.78, 0.99)  0.89 (0.80, 0.99)  0.80 (0.63, 1.01) | 0.04  0.04  0.07 | 1.19 (1.04, 1.36)  1.29 (1.15, 1.45)  1.29 (0.86, 1.94) | 0.01  <0.001  0.21 | 1.06 (0.96, 1.17)  1.10 (0.99, 1.22)  1.13 (0.93, 1.38) | 0.23  0.07  0.21 |
| Pharmacy trainee present* | 1.01 (0.93, 1.08) | 0.86 | 1.00 (0.93, 1.07) | 0.94 | 1.03 (0.97, 1.09) | 0.39 |
| Nurse-to-patient ratio* (reference: 1:2)  ≤1  1-2  >2 | 0.78 (0.67, 0.90)  0.79 (0.67, 0.94)  1.08 (0.86, 1.35) | <0.001  0.006  0.5 | 0.81 (0.67, 0.99)  0.84 (0.64, 1.10)  1.49 (1.10, 2.00) | 0.04  0.21  0.009 | 0.82 (0.70, 0.96)  0.97 (0.58, 1.62)  1.26 (1.04, 1.51) | 0.01  0.91  0.02 |
| Percent of assigned ICU teams pharmacist rounded with**§*** | 1.00 (1.00, 1.00) | 0.09 | 1.00 (1.00, 1.00) | 0.08 | 1.00 (1.00, 1.00) | 0.03 |

APP: advanced practice provider, CI: confidence interval, CMM: comprehensive medication management, CMS: Center for Medicare & Medicaid Services, HDA: hazard of discharge alive, ICU: intensive care unit, MRC-ICU: medication regimen complexity- intensive care unit, OR: odds ratio, SOFA: sequential organ failure assessment, VA: Veterans Affairs

*indicates that this is reflective of only the value on the first day of ICU stay

**†**Inclusion criteria specified ≥18 years of age; however, some institutions allowed patients ≥18 years of age in the pediatric ICU

**‡**CMM delivered on interprofessional rounds: the pharmacist attended interprofessional rounds and verbally provided CMM (including review of medications and recommendations); CMM delivered not on interprofessional rounds: the pharmacist provided CMM (reviewed medications and provided recommendations) but did not attend interprofessional rounds; Abbreviated CMM delivered: the pharmacist provided abbreviated CMM which includes brief medication review but does not include full patient review (e.g. progress notes, results) and recommendations were provided outside of rounds; CMM not delivered (absence of CMM): the patient received no medication review from a pharmacist outside of pharmacokinetic monitoring and prospective medication order verification

**§**Percent of assigned ICU teams pharmacist rounded is calculated by total number of ICU medical teams the pharmacist attended and provided CMM on interprofessional rounds for divided by total number of ICU medical teams providing care for patients that the pharmacist was assigned to provide CMM for

This Table examines likelihood of discharge, and odds ratios under 1 indicate decreased likelihood of discharge, whereas odds ratios greater than 1 indicate increased likelihood of discharge.
