## Supplementary material for "Impact of Early Interprofessional Involvement with a Critical Care Pharmacist on Patient Outcomes in the Intensive Care Unit": Table 1

**Table 1. Pharmacist and Institution Characteristics**

| **Institution Characteristics (N=64)** | | **Pharmacist Characteristics (N=216)**** | |
| --- | --- | --- | --- |
| **Variable** |  | **Variable** |  |
| **Hospital Type** | | **Degree(s) earned** | |
| Academic Medical Center | 39 (60.9) | Doctor of Pharmacy | 213 (99.1) |
| Community Teaching Hospital | 16 (25.0) | Bachelor of Science in Pharmacy | 24 (11.2) |
| Community Non-Teaching Hospital | 8 (12.5) | Master of Science | 16 (7.4) |
| Government/VA/Military | 1 (1.6) | Doctor of Philosophy | 1 (0.5) |
| **Hospital Beds** | | Other | 55 (25.6) |
| <200 | 2 (3.1) | **Highest level of postgraduate training** | |
| 200-400 | 17 (26.6) | Post-graduate Year 1 | 49 (22.8) |
| 401-600 | 14 (21.9) | Post-graduate Year 2 | 144 (67.0) |
| >600 | 31 (48.4) | Fellowship | 4 (1.9) |
| **ICU Beds** | | No postgraduate training | 18 (8.4) |
| < 26 | 6 (9.4) | **Active board certification** | |
| 26-50 | 13 (20.3) | BCCCP | 155 (71.8) |
| 51-75 | 9 (14.1) | BCPS | 79 (36.6) |
| 76-100 | 6 (9.4) | Board Certification in Other Specialties | 20 (9.3) |
| 101-150 | 10 (15.6) | No active board certification | 19 (8.8) |
| >150 | 20 (31.3) | **Years since graduation from training** | |
| **Region** | | Years since graduation from highest training program | 8.3 (±7.1) |
| Northeast | 11 (17.2) | **Type of ICU most commonly practiced in** | |
| Southeast | 20 (31.3) | Medical | 59 (27.3) |
| Midwest | 16 (25.0) | Surgical/Trauma | 35 (16.2) |
| Northwest | 4 (6.3) | Mixed Medical/Surgical | 34 (15.7) |
| Southwest | 11 (17.2) | Cardiothoracic/Cardiac | 27 (12.5) |
| International (non-USA) | 2 (3.1) | Neurosurgery/Neurology | 14 (6.5) |
| **Adult Trauma Center** | | Other | 30 (13.9) |
| Level I | 33 (51.6) | Not specified | 17 (7.9) |
| Level II | 14 (21.9) |  |  |
| Level III | 6 (9.4) |  |  |
| Level IV | 1 (1.6) |  |  |
| N/A | 10 (15.6) |  |  |
| **Hospital CMS star rating** |  |  |  |
| 2 | 12 (20.3) |  |  |
| 3 | 34 (57.6) |  |  |
| 4 | 11 (18.6) |  |  |
| 5 | 2 (3.4) |  |  |
| Missing/N/A* | 5 (7.8) |  |  |
| **Pharmacist-to-team ratio on a weekday, 1:X** |  |  |  |
| Pharmacist-to-team ratio on a weekday, 1:X | 2.1 (±1.6) |  |  |
| Missing | 2 (3.1) |  |  |
| **Float FTEs** |  |  |  |
| Total adult critical care float FTEs employed by the institution | 1.5 (±3.2) |  |  |
| **Cross Coverage Approach (how are patients covered if a pharmacist is on PTO)** |  |  |  |
| Float pharmacist covers that unit | 29 (46.8) |  |  |
| A specific critical care pharmacist would cover their regular patients plus the person on PTO’s patients | 15 (24.2) |  |  |
| Multiple pharmacists would split up the unit to cover their own patients plus a portion of the unit where the pharmacist is on PTO | 2 (3.2) |  |  |
| A centralized pharmacist would cover that unit | 2 (3.2) |  |  |
| Other | 14 (22.6) |  |  |
| Missing | 2 (3.1) |  |  |

BCPS: Board Certified Pharmacotherapy Specialist; BCCCP: Board Certified Critical Care Pharmacist; CMS: Center for Medicare & Medicaid Services, FTE: full-time equivalent (1.0 = 40 hours per week), ICU: Intensive Care Unit, PTO: paid time off

Data reported as n (%) or mean (±SD) unless otherwise specified

*2 centers do not have a CMS star rating as they are located outside of the United States (US). 2 US hospitals included also did not have a CMS star rating per the publicly available CMS website as of April 2026.

**Missing demographic data from 1 pharmacist
