## Supplementary material for "Impact of Early Interprofessional Involvement with a Critical Care Pharmacist on Patient Outcomes in the Intensive Care Unit": Table 2

**Table 2. Patient demographics and outcomes by ICU pharmacist coverage in the first 24 hours**

| **Variable** | **Overall (21,835)** | **CMM delivered on interprofessi-onal rounds (16,605)** | **CMM delivered not on interprofessi-onal rounds (2,998)** | **Abbreviated CMM delivered (810)** | **CMM not delivered (absence of CMM) (1,321)** | **Unknown/ Missing (101)** | **p-value** |
| --- | --- | --- | --- | --- | --- | --- | --- |
| ***Secondary Variable*** | | | | | | | |
| Pharmacist-to-patient ratio (ICU patients)*  Missing | 17.7 (±10.5)  1148 (5.3) | 15.9 (±6.9)  124 (0.7) | 21.2 (±11.5)  65 (2.2) | 43.3 (±23.8)  75 (9.3) | 18.5 (±16.3)  882 (66.8) | 19.7 (±8.7)  2 (2.0) | <0.001 |
| ***Co-variates*** | | | | | | | |
| Age, years | 61.6 (±16.7) | 61.5 (±16.7) | 62.0 (±16.7) | 61.1 (±16.5) | 62.2 (±16.7) | 58.6 (±18.1) | 0.1 |
| Sex, female | 9226 (42.3) | 7031 (42.3) | 1264 (42.2) | 351 (43.3) | 546 (41.3) | 34 (33.7) | 0.8 |
| SOFA Score*  Missing | 5.2 (±4.1)  25 (0.1) | 5.2 (±4.1)  22 (0.1) | 4.9 (±4.0)  3 (0.1) | 5.3 (±4.2)  0 | 5.1 (±4.1)  0 | 6.5 (±4.4)  0 | <0.001 |
| MRC-ICU Score*  Missing | 11.4 (±6.6)  2 (<0.1) | 11.5 (±6.7)  1 (<0.1) | 11.1 (±6.5)  0 | 11.3 (±6.7)  0 | 11.2 (±6.2)  1 (<0.1) | 13.7 (±6.5)  0 | <0.001 |
| ICU admission day of the week  Monday  Tuesday  Wednesday  Thursday  Friday | 4634 (21.2)  4599 (21.1) 4364 (20.0)  4386 (20.1)  3852 (17.6) | 3,891 (23.4)  3,923 (23.6)  3,653 (22.0)  3,586 (21.6)  1,552 (9.3) | 570 (19.0)  573 (19.1)  536 (17.9)  560 (18.7)  759 (25.3) | 42 (5.2)  24 (3.0)  52 (6.4)  52 (6.4)  640 (79.0) | 118 (8.9)  64 (4.8)  101 (7.6)  166 (12.6)  872 (66.0) | 13 (12.9)  15 (14.9)  22 (21.8)  22 (21.8)  29 (28.7) | <0.001 |
| ICU type**†***  Medical  Surgical/Trauma  Surgical  Cardiothoracic  Cardiac  Neurosurgery/Neurology  Mixed Medical-Surgical  Pediatric  Burn  Other  Missing | 6568 (30.1)  2757 (12.6)  2026 (9.3)  2272 (10.4)  1964 (9.0)  2928 (13.4)  3113 (14.3)  27 (0.1)  141 (0.6)  37 (0.2)  2 (<0.1) | 5153 (31.0)  2118 (12.8)  1456 (8.8)  1744 (10.5)  1440 (8.7)  2116 (12.7)  2433 (14.7)  23 (0.1)  88 (0.5)  32 (0.2)  2 (<0.1) | 791 (26.4)  256 (8.5)  333 (11.1)  329 (11.0)  325 (10.8)  533 (17.8)  402 (13.4)  2 (0.1)  25 (0.8)  2 (0.1)  0 | 310 (38.3)  137 (16.9)  81 (10.0)  61 (7.5)  75 (9.3)  93 (11.5)  42 (5.2)  0  11 (1.4)  0  0 | 282 (21.3)  225 (17.0)  135 (10.2)  130 (9.8)  122 (9.2)  174 (13.2)  234 (17.7)  2 (0.2)  14 (1.1)  3 (0.2)  0 | 32 (31.7)  21 (20.8)  21 (20.8)  8 (7.9)  2 (2.0)  12 (11.9)  2 (2.0)  0  3 (3.0)  0  0 | <0.001 |
| Hospital type  Academic Medical Center  Community - Teaching  Community - Non-teaching  Government/VA/Military | 14590 (66.8)  5322 (24.4)  1797 (8.2)  126 (0.6) | 11055 (66.6)  4184 (25.2)  1297 (7.8)  69 (0.4) | 2022 (67.4)  596 (19.9)  325 (10.8)  55 (1.8) | 640 (79.0)  142 (17.5)  26 (3.2)  2 (0.2) | 795 (60.2)  377 (28.5)  149 (11.3)  0 | 78 (77.2)  23 (22.8)  0  0 | <0.001 |
| Medical team coverage*  Attending physician only  Attending physician and APP  Attending physician and medical residents/fellows  Attending physician and medical residents/fellows and APP  Missing | 1772 (8.2)  5065 (23.6)  7178 (33.4)  7464 (34.8)  356 (1.6) | 1183 (7.2)  3798 (23.1)  5722 (34.8)  5721 (34.8)  181 (1.1) | 435 (14.8)  733 (25.0)  703 (23.9)  1065 (36.3)  62 (2.1) | 32 (4.2)  190 (24.6)  264 (34.2)  285 (37.0)  39 (4.8) | 122 (9.8)  335 (26.8)  450 (36.0)  342 (27.4)  72 (4.8) | 0  9 (9.1)  39 (39.4)  51 (51.5)  2 (2.0) | <0.001 |
| Hospital CMS star rating  2  3  4  5  Missing | 4853 (22.8)  11205 (52.6)  4922 (23.1)  334 (1.6)  521 (2.4) | 3369 (20.9)  8433 (52.2)  4014 (24.9)  334 (2.1)  455 (2.7) | 958 (32.1)  1429 (47.9)  597 (20.0)  0  14 (0.5) | 192 (24.4)  502 (63.7)  94 (11.9)  0  22 (2.7) | 327 (25.3)  789 (61.1)  175 (13.6)  0  30 (2.3) | 7 (6.9)  52 (51.5)  42 (41.6)  0  0 | <0.001 |
| Pharmacy trainee present*  Missing | 9041 (42.2)  406 (1.9) | 7518 (45.9)  213 (1.3) | 952 (32.0)  23 (0.8) | 302 (38.5)  26 (3.2) | 234 (19.8)  141 (10.7) | 35 (35.7)  3 (3.0) | <0.001 |
| Nurse-to-patient ratio, 1:X*  ≤1  1-2  2  >2  Missing | 2432 (12.0)  47 (0.2)  17620 (86.7)  229 (1.1)  1507 (6.9) | 2053 (12.9)  27 (0.2)  13,643 (85.9)  158 (1.0)  724 (4.4) | 230 (8.6)  13 (0.5)  2405 (89.4)  41 (1.5)  309 (10.3) | 26 (3.7)  1 (0.1)  663 (94.8)  9 (1.3)  111 (13.7) | 112 (11.6)  0  834 (86.2)  21 (2.2)  354 (26.8) | 11 (12.0)  6 (6.5)  75 (81.5)  0  9 (8.9) | <0.001 |
| Percent of assigned ICU teams pharmacist rounded with**§***  Missing | 66.2 (±34.0)  1157 (5.3) | 74.1 (±29.5)  128 (0.8) | 36.1 (±29.6)  65 (2.2) | 11.4 (±24.2)  75 (9.3) | 64.4 (±36.5)  887 (67.1) | 58.3 (±33.0)  2 (2.0) | <0.001 |
| CMM delivered every day of ICU stay | 16696 (76.5) | 13498 (81.3) | 2431 (81.1) | 690 (85.2) | 0 (0.0) | 77 (76.2) | <0.001 |
| Percent days, only attending physician**⁂**  Missing | 8.6 (±26.4)  1326 (6.1) | 7.9 (±25.3)  965 (5.8) | 14.3 (±33.2)  192 (6.4) | 3.4 (±16.7)  55 (6.8) | 8.8 (±26.5)  106 (8.0) | 0.4 (±4.0)  8 (7.9) | <0.001 |
| Required dialysis during ICU stay  Missing | 2560 (11.7)  2 (<0.1) | 1937 (11.7)  0 | 346 (11.5)  0 | 98 (12.1)  0 | 158 (12.0)  2 (0.2) | 21 (20.8)  0 | 0.079 |
| Required mechanical ventilation | 10733 (49.2) | 8197 (49.4) | 1384 (46.2) | 396 (48.9) | 688 (52.1) | 68 (67.3) | <0.001 |
| Mean pharmacist-to-patient ratio (ICU Patients) across hospital stay  Missing | 19.4 (±10.1)  279 (1.3) | 18.3 (±8.9)  19 (0.1) | 22.7 (±11.1)  23 (0.8) | 32.7 (±17.1)  20 (2.5) | 16.9 (±9.4)  217 (16.4) | 20.9 (±7.5)  0 | <0.001 |
| Mean pharmacist-to-patient ratio (all patients) across hospital stay  Missing | 27.2 (±22.7)  279 (1.3) | 25.2 (±20.1)  19 (0.1) | 33.0 (±25.5)  23 (0.8) | 54.0 (±43.2)  20 (2.5) | 21.5 (±14.0)  217 (16.4) | 26.8 (±11.8)  0 | <0.001 |
| ***Outcomes*** | | | | | | | |
| In hospital mortality | 3105 (14.2) | 2394 (14.4) | 377 (12.6) | 100 (12.3) | 214 (16.2) | 20 (19.8) | 0.003 |
| Hospital LOS, days  Missing | 12.2 (±16.9)  2 (<0.1) | 11.9 (±16.3)  2 (<0.1) | 12.2 (±15.7)  0 | 13.6 (±16.8)  0 | 14.9 (±23.4)  0 | 25.2 (±27.7)  0 | <0.001 |
| ICU LOS, days | 5.7 (±9.7) | 5.5 (±9.2) | 5.5 (±8.1) | 6.2 (±8.4) | 7.7 (±15.8) | 15.9 (±23.8) | <0.001 |
| Duration of mechanical ventilation, days  Missing | 2.3 (±20.5)  1 (<0.1) | 2.1 (±5.6)  1 (<0.1) | 2.9 (±53.6)  0 | 2.1 (±5.2)  0 | 2.8 (±6.6)  0 | 5.8 (±9.0)  0 | <0.001 |

Data presented as mean (±SD) or n (%)

APP: advanced practice provider, CMM: comprehensive medication management, CMS: Center for Medicare & Medicaid Services, ICU: intensive care unit, LOS: length of stay, MRC-ICU: medication regimen complexity- intensive care unit, SOFA: sequential organ failure assessment, VA: Veterans Affairs

*indicates that this is reflective of only the value on the first day of ICU stay

**†**Inclusion criteria specified ≥18 years of age; however, some institutions allowed patients ≥18 years of age in the pediatric ICU

**‡**CMM delivered on interprofessional rounds: the pharmacist attended interprofessional rounds and verbally provided CMM (including review of medications and recommendations); CMM delivered not on interprofessional rounds: the pharmacist provided CMM (reviewed medications and provided recommendations) but did not attend interprofessional rounds; Abbreviated CMM delivered: the pharmacist provided abbreviated CMM which includes brief medication review but does not include full patient review (e.g. progress notes, results) and recommendations were provided outside of rounds; CMM not delivered (absence of CMM): the patient received no medication review from a pharmacist outside of pharmacokinetic monitoring and prospective medication order verification

**§**Percent of assigned ICU teams pharmacist rounded is calculated by total number of ICU medical teams the pharmacist attended and provided CMM on interprofessional rounds for divided by total number of ICU medical teams providing care for patients that the pharmacist was assigned to provide CMM for
