## Supplementary material for "Impact of Early Interprofessional Involvement with a Critical Care Pharmacist on Patient Outcomes in the Intensive Care Unit": Table 3

**Table 3. Unadjusted and adjusted odds ratios for GEE model for primary outcome of mortality**

| **Variable** | **Univariable** | | **Multivariable** | |
| --- | --- | --- | --- | --- |
|  | **Odds ratio (CI)** | **P-value** | **Odds Ratio (CI)** | **P-value** |
| ***Primary Variable*** | | | |  |
| ICU pharmacist coverage 1st 24 hours‡* (reference: CMM delivered on interprofessional rounds)  CMM delivered not on interprofessional rounds  Abbreviated CMM delivered  CMM not delivered (absence of CMM)  Unknown | 0.90 (0.78-1.05)  0.91 (0.80-1.05)  1.13 (0.96-1.32)  1.64 (0.99-2.72) | 0.19  0.2  0.14  0.06 | 1.01 (0.88-1.16)  0.92 (0.71-1.21)  1.23 (1.04-1.46)  1.30 (0.70-2.44) | 0.85  0.56  0.02  0.41 |
| ***Secondary Variable*** | | | | |
| Pharmacist-to-patient ratio (ICU patients)* | 1.00 (1.00-1.01) | 0.76 | 1.00 (1.00-1.01) | 0.17 |
| ***Co-variates*** | | | | |
| Age, years | 1.02 (1.02-1.02) | <0.001 | 1.03 (1.02-1.03) | <0.001 |
| Sex, female | 0.97 (0.90-1.05) | 0.47 | 1.01 (0.92-1.11) | 0.81 |
| SOFA Score* | 1.25 (1.23-1.28) | <0.001 | 1.31 (1.27-1.35) | <0.001 |
| MRC-ICU Score* | 1.08 (1.07-1.09) | <0.001 | 1.04 (1.02-1.05) | <0.001 |
| ICU admission day of the week (reference: Monday)  Tuesday  Wednesday  Thursday  Friday | 0.96 (0.85-1.07)  0.97 (0.86-1.10)  0.98 (0.86-1.10)  1.01 (0.89-1.13) | 0.45  0.65  0.68  0.93 | 0.98 (0.87-1.11)  0.95 (0.84-1.07)  0.99 (0.87-1.12)  0.94 (0.82-1.07) | 0.78  0.4  0.86  0.36 |
| ICU type†* (reference: medical ICU)  Surgical/Trauma  Surgical  Cardiothoracic  Cardiac  Neurosurgery/Neurology  Mixed Medical-Surgical  Pediatric  Burn  Other | 0.54 (0.44-0.67)  0.50 (0.37-0.67)  0.32 (0.23-0.45)  0.73 (0.59-0.90)  0.52 (0.43-0.63)  0.72 (0.57-0.91)  0.16 (0.03-0.83)  0.34 (0.18-0.63)  0.82 (0.47-1.44) | <0.001  <0.001  <0.001  0.004  <0.001  0.007  0.03  <0.001  0.5 | 0.77 (0.63-0.95)  0.51 (0.38-0.69)  0.22 (0.14-0.35)  0.93 (0.72-1.20)  1.16 (0.94-1.43)  0.75 (0.61-0.93)  0.67 (0.18-2.50)  0.64 (0.36-1.14)  1.18 (0.85-1.64) | 0.01  <0.001  <0.001  0.58  0.17  0.008  0.55  0.13  0.33 |
| Hospital type (reference: academic medical center)  Community - Teaching  Community - Non-teaching  Government/VA/Military | 0.97 (0.74-1.26)  0.88 (0.63-1.24)  0.73 (0.61-0.86) | 0.82  0.47  <0.001 | 0.95 (0.76-1.20)  0.79 (0.62-1.02)  0.95 (0.75-1.19) | 0.68  0.07  0.66 |
| Medical team coverage* (reference: only attending physician)  Attending physician and APP  Attending physician and medical residents/fellows  Attending physician and medical residents/fellows and APP | 0.93 (0.69-1.26)  1.23 (0.88-1.73)  1.08 (0.76-1.54) | 0.65  0.23  0.67 | 0.78 (0.62-0.98)  0.89 (0.66-1.19)  0.90 (0.67-1.21) | 0.03  0.43  0.49 |
| Hospital CMS star rating (reference: 3)  2  4  5 | 0.93 (0.73-1.19)  1.06 (0.80-1.42)  0.80 (0.41-1.56) | 0.56  0.67  0.51 | 1.03 (0.84-1.28)  0.96 (0.74-1.23)  0.68 (0.43-1.08) | 0.75  0.73  0.1 |
| Pharmacy trainee present* | 1.01 (0.93-1.10) | 0.79 | 1.01 (0.91-1.11) | 0.92 |
| Nurse-to-patient ratio* (reference: 1:2)  ≤1  1-2  >2 | 1.79 (1.39-2.32)  0.89 (0.57-1.39)  0.60 (0.43-0.83) | <0.001  0.60  0.002 | 1.39 (1.03-1.88)  0.94 (0.60-1.46)  0.76 (0.56-1.04) | 0.03  0.77  0.09 |
| Percent of assigned ICU teams pharmacist rounded with**§*** | 1.00 (1.00-1.00) | 0.11 | 1.00 (1.00-1.00) | 0.08 |
| SOFA-MRC-ICU interaction term | 1.01 (1.01-1.01) | <0.001 | 1.00 (1.00-1.00) | <0.001 |

APP: advanced practice provider, CI: confidence interval, CMM: comprehensive medication management, CMS: Center for Medicare & Medicaid Services, ICU: intensive care unit, LOS: length of stay, MRC-ICU: medication regimen complexity- intensive care unit, SOFA: sequential organ failure assessment, VA: Veterans Affairs

*indicates that this is reflective of only the value on the first day of ICU stay

**†**Inclusion criteria specified ≥18 years of age; however, some institutions allowed patients ≥18 years of age in the pediatric ICU

**‡**CMM delivered on interprofessional rounds: the pharmacist attended interprofessional rounds and verbally provided CMM (including review of medications and recommendations); CMM delivered not on interprofessional rounds: the pharmacist provided CMM (reviewed medications and provided recommendations) but did not attend interprofessional rounds; Abbreviated CMM delivered: the pharmacist provided abbreviated CMM which includes brief medication review but does not include full patient review (e.g. progress notes, results) and recommendations were provided outside of rounds; CMM not delivered (absence of CMM): the patient received no medication review from a pharmacist outside of pharmacokinetic monitoring and prospective medication order verification

**§**Percent of assigned ICU teams pharmacist rounded is calculated by total number of ICU medical teams the pharmacist attended and provided CMM on interprofessional rounds for divided by total number of ICU medical teams providing care for patients that the pharmacist was assigned to provide CMM for
